# Anorexia nervosa is associated with higher brain mu-opioid receptor availability

**DOI:** 10.1101/2024.03.26.24304878

**Authors:** Kyoungjune Pak, Jouni Tuisku, Henry K. Karlsson, Jussi Hirvonen, Eleni Rebelos, Laura Pekkarinen, Lihua Sun, Aino Latva-Rasku, Semi Helin, Johan Rajander, Max Karukivi, Pirjo Nuutila, Lauri Nummenmaa

## Abstract

Anorexia nervosa (AN) is a severe psychiatric disorder, characterized by restricted eating, fear to gain weight, and a distorted body image. Mu-opioid receptor (MOR) functions as a part of complex opioid system and supports both homeostatic and hedonic control of eating behavior. Thirteen patients with AN and thirteen healthy controls (HC) were included in this study. We measured 1) MOR availability with [^11^C]carfentanil positron emission tomography (PET), 2) brain glucose uptake (BGU) with 2-deoxy-2[^18^F]fluoro-D-glucose ([^18^F]FDG) PET during hyperinsulinemic-euglycemic clamp and 3) blood-oxygen-level-dependent signal with functional magnetic resonance imaging. All subjects underwent a screening visit consisting of physical examination, anthropometric measurements, fasting blood samples, an oral glucose tolerance test, psychiatric assessment, and an inquiry regarding medical history. Body fat mass (%) was measured and M value was calculated. MOR availability from caudate and putamen was higher in patients with AN and those from nucleus accumbens (NAcc) and thalamus showed the higher trend in patients with AN. There was no area where MOR availability was lower in patients with AN. BGU was not different between AN and HC. MOR availability and BGU were negatively correlated in caudate, NAcc and thalamus and showed the trend of negative association in putamen. In conclusion, AN is associated with higher MOR availability in the brain regions implicated in reward processing, while BGU remains unaltered. Therefore, the endogenous opioid system might be one of the key components underlying AN. This better understanding of AN could support the development of new treatments for AN.

## INTRODUCTION

Anorexia nervosa (AN) is a severe psychiatric disorder, characterized by restricted eating, fear to gain the weight, and a distorted body image ^1^. The peak age of AN onset is during adolescence, and more than 90% of AN patients are females ^1^. The exact cause of AN is unknown, however, a combination of genetic, neurobiological, psychological and environmental factors might affect the development of AN ^2^. Comorbidities of mood disorders, anxiety disorders, obsessive-compulsive disorders, autism spectrum disorder, and attention-deficit hyperactivity disorder are common in AN ^3^. Also, patients with AN can have medical complications as a direct result of weight loss and malnutrition, such as hypoglycemia, osteoporosis, sarcopenia, cerebral atrophy, organ damage, and sudden death from arrhythmia ^4^.

Treatment of AN aims to restore body weight, mend the body image, eliminate the problematic eating patterns and develop long-term behavioral changes ^3^. Current therapeutic approaches of AN include psychotherapy, medications as well as dietary advice ^3^. The prognosis of patients with AN remains however poor, with the highest mortality rate among all psychiatric disorders ^1^. Among the surviving patients, less than 50% recover, whereas one third improve, and 20% remain chronically ill ^5^. Therefore, understanding the pathophysiology of AN is crucial in the development of an effective treatment for AN.

Alterations of multiple neurotransmitter systems are linked with AN. Positron emission tomography (PET) using receptor or transporter binding radiopharmaceuticals has been utilized to understand the role of neurotransmitter systems in AN. Increased serotonin 1A receptor of temporal, frontal lobes and amygdala ^6^, decreased serotonin 2A receptor of frontal, parietal and occipital lobes ^7^ and increased cannabinoid 1 receptor of both cortical and subcortical areas ^8^ was reported with a regard to reward processing in eating behavior. However, there was no significant difference of dopamine receptor between patients with AN and healthy controls (HC) ^9^, although mesolimbic pathway in AN works differently during reward learning and responding ^10^. Also, fasting brain glucose uptake (BGU) was investigated with 2-deoxy-2[^18^F]fluoro-D-glucose ([^18^F]FDG) PET showing the inconsistent results of either global hypometabolism ^11^ or hypermetabolism of frontal lobe, hippocampus, amygdala and insula in AN ^12^.

The opioid system modulates motivation and reward processing and consequently is intimately linked with appetite and weight regulation ^13^. Opioid agonists increase and opioid receptor antagonists decrease food intake and opioid receptor agonists stimulate food intake and the hedonic responses to palatable foods ^14^. Feeding also leads to the release of endogenous opioid in humans ^15^. In morbid obesity, mu-opioid receptor (MOR) availability was decreased ^13^ and weight loss after bariatric surgery normalizes MOR availability, suggesting weight-dependent regulation of MORs ^16^. Also, lower MOR availability is associated with the increased familial obesity risk such as parental obesity or diabetes ^17^. Therefore, MOR functions as a part of complex opioid system and supports both homeostatic and hedonic control of eating behavior ^18^.

Here, we investigated whether MOR and BGU are altered in patients with AN with comparison to HC. We used PET scans with MOR-specific ligand [^11^C]carfentanil to quantify MOR availability and with [^18^F]FDG during hyperinsulinemic, euglycemic clamp, to study brain glucose metabolism. We hypothesized that AN would be associated with the alteration of MOR with a regard to reward processing and BGU with a regard to insulin sensitivity.

## MATERIALS AND METHODS

The study was conducted in accordance with the Declaration of Helsinki and approved by the Ethics Committee of the Hospital District of Southwest Finland. This study is a part of AVAIN project registered at ClinicalTrials.gov (Anorexia Nervosa and Its Effects on Brain Function, Body Metabolism and Their Interaction, NCT05101538). All participants signed ethics committee-approved informed consent forms prior to inclusion.

### Subjects and study design

We recruited 13 patients with AN for the study. All patients with AN were required to meet the following criteria: 1) female, 2) age 18-32 years, 3) body mass index (BMI) < 17.5kg/m^2^, 4) currently fulfilling modified DSM-IV diagnosis of AN with or without amenorrhea, 5) AN onset before the age of 25 years, 6) no lifetime history of binge eating and 7) diagnosed less than 2 years ago. Patients with AN were compared with 13 HC who met the following criteria: 1) female, 2) age 18-32 years, 3) BMI 20-25 kg/m^2^ and 4) no lifetime history of obesity (BMI ≥ 30kg/m^2^) or eating disorders. Exclusion criteria were any chronic disease or medication that would affect glucose metabolism or neurotransmission, history of psychiatric disorders (apart from AN), and abusive use of alcohol. Subjects underwent a screening consisting of physical examination, anthropometric measurements, fasting blood samples, a 75 g oral glucose tolerance test (OGTT), psychiatric assessment, and an inquiry regarding medical history. Body fat mass (%) was measured with an air displacement plethysmograph (the Bod Pod system, software version 5.4.0, COSMED, Inc., Concord, CA, USA) after at least 4 h of fasting. M value was calculated as a measure of insulin sensitivity as previously described ^19^ and expressed as per kilogram of fat-free mass, because this normalization minimizes differences due to sex, age, and body weight ^20^.

### Brain PET acquisition

We measured MOR availability with high-affinity agonist radioligand [^11^C]carfentanil with high sensitivity, and specificity for MORs. BGU was quantified with radioligand [^18^F]FDG during hyperinsulinemic-euglycemic clamp. Scans were done on two separate visits. The subjects were advised to abstain from physical exercise in the PET scan days and the day before. PET studies were done after a 12-h overnight fast. Computed tomography scans were acquired before PET scans for attenuation correction. For [^11^C]carfentanil PET scan, a catheter was placed in the subject’s left antecubital vein for tracer administration. After injection of [^11^C]carfentanil, PET scan was acquired for 51 min (13 frames). For [^18^F]FDG PET scan, hyperinsulinemic-euglycemic clamp was applied as previously described ^21, 22^ to measure whole-body insulin sensitivity. After reaching steady glycemia, [^18^F]FDG was injected intravenously and a peripheral scan for heart (5 min) and abdomen (25 min) was done before brain dynamic PET scan for 15 mins (3 frames*5 min). Arterialized venous blood samples were taken to measure plasma activity, glucose, and insulin. The plasma radioactivity was measured by automatic gamma-counter (Wizard 1480 3”, Wallac, Turku, Finland). Anatomical T1-weighted magnetic resonance (MR) images (TR, 8.1 ms; TE, 3.7 ms; flip angle, 7°; scan time, 263 s; 1 mm^3^ isotropic voxels) were obtained for anatomical normalization and reference; T2-weighted and fluid-attenuated inversion recovery (FLAIR) images were obtained to exclude significant brain pathology.

### Brain PET analysis

An in-house automated processing tool (Magia ^23^; https://github.com/tkkarjal/magia) was used to process the PET data. Processing began with motion-correction of the PET data followed by coregistration of the PET and MRI images. Magia uses FreeSurfer (http://surfer.nmr.mgh.harvard.edu/) to define the regions of interest (ROIs) as well as the reference regions. The regional (ROI-wise) kinetic modeling was based on extracting ROI-wise time-activity curves. Voxel-wise parametric images were also generated, spatially normalized to MNI-space and finally smoothed using a Gaussian kernel. BGU-estimates (μmol/min/100g) obtained from FDG PET data are based on fractional uptake rate multiplied by the average plasma glucose concentration from the injection until the end of the brain scan, divided by the lumped constant for the brain (set at 0.65) ^24^. MOR availability was quantified by binding potential (BP_ND_), which is the ratio of specific binding to non-displaceable binding in the tissue with simplified reference tissue model. Occipital cortex was used as the reference region ^25^. For a full-volume comparison between AN and HC, the statistical threshold in statistical parametric mapping analysis was set at a cluster level and corrected with false discovery rate with p < 0.05.

### fMRI acquisition

fMRI scans were acquired after overnight fasting. We used a previously established task protocol ^26^ for inducing anticipatory reward, by showing the participants pictures of palatable (for example, chocolate, pizza, cakes) and bland (for example, lentils, cereal, eggs) food pictures. This task simulates situations where appetite is triggered by anticipating the actual feeding via visual food cues, such as those in advertisements. The pictures were rated in a previous study by independent participants; the ratings showed that the appetizing foods were evaluated more pleasant than the bland foods, t(31) = 4.67, p < 0.001 ^27^. During the tasks, functional data were acquired with gradient echo-planar imaging (EPI) sequence, sensitive to the blood-oxygen level-dependent imaging (BOLD) signal. Participants viewed alternating 16.2-s epochs with pictures of palatable or non-palatable foods. Each epoch contained nine stimuli from one category intermixed with fixation crosses. Each food stimulus was presented on either the right or the left side of the screen, and participants were instructed to indicate its location by pressing corresponding buttons. This task was used simply to ensure that participants had to pay attention to the stimuli. Stimulus delivery was controlled by the Presentation software (Neurobehavioral System, Inc.). Functional data were acquired using 3-Tesla Philips Ingenuity PET/MR scanner and using EPI sequence with the following parameters: TR, 2,600 ms; TE, 30 ms; flip angle 75°, 240*240*135 mm^3^ FOV, 3*3*3 mm^3^ voxel size. Each volume consisted of 45 interleaved axial slices acquired in ascending order. A total of 165 functional volumes were acquired, with additional five dummy volumes acquired and discarded at the beginning of each run. Anatomical reference images were acquired using a T1-weighted sequence with following parameters: TR 8.1 ms, TE 3.7 ms, flip angle 7°, 256*56*176 mm^3^ FOV, 1*1*1 mm^3^ voxel size.

### fMRI analysis

Preprocessing of MRI images was performed using FMRIPREP (v.1.3.0.2) ^28^, a Nipype (v.1.1.9) ^29^ based tool. Each T1-weighted image was corrected for intensity non-uniformity with N4BiasFieldCorrection (v.2.1.0) ^30^ and skull-stripped on the basis of the OASIS template with antsBrainExtraction. sh workflow (ANTs v.2.2.0). Brain surfaces were reconstructed using recon-all from FreeSurfer (v.6.0.0) ^31^, and the brain mask estimated previously was refined with a custom variation of the method to reconcile ANTs-and FreeSurfer-derived segmentations of the cortical grey matter using Mindboggle ^32^. Spatial normalization to the ICBM 152 non-linear asymmetrical template v.2009c ^33^ was done with non-linear registration using the antsRegistration tool ^34^. Tissue segmentation of cerebrospinal fluid, white matter and grey matter was performed on the brain-extracted T1w using fast ^35^ from FSL (v.5.0.9). fMRI images were slice time corrected with 3dTshift from AFNI (v.16.2.07) ^36^ and motion corrected with mcflirt ^37^ from FSL. This was followed by coregistration to the corresponding T1w via boundary-based registration ^38^ with nine degrees of freedom, using bbregister from FreeSurfer. These steps were concatenated and applied in a single step via antsApplyTransforms with the Lanczos interpolation. Physiological noise regressors were obtained using CompCor ^39^, where principal components were estimated for temporal and anatomical variants. A mask to exclude signal with cortical origin was created via eroding the brain mask, ensuring it merely originating from subcortical structures. Six temporal CompCor components were then calculated including the top 5% variable voxels within that subcortical mask. For anatomical CompCor components, six components were calculated within the intersection of the subcortical mask and the union of cerebral spinal fluid and white matter masks, after projection to the native space of each functional run. Frame-wise displacement ^40^ was calculated for each functional run using the implementation of Nipype. Finally, ICA-based automatic removal of motion artefacts was used to generate aggressive noise regressors and also to create a variant of data that is non-aggressively denoised ^41^. The quality of images was assessed using FMRIPREP’s visual reports, where data were manually inspected for whole-brain field of view coverage, alignment to the anatomical images and artefacts.

### Statistical analysis

Data are presented as mean±SD. Normality of data distribution was tested with Shapiro-Wilk test. Two-sample t-test was used to compare MOR availability, BGU, and subjects’ characteristics across groups. Pearson correlation analysis was used to determine the association between MOR availability, BGU and BMI, M value. Statistical analysis was carried out in R Statistical Software (The R Foundation for Statistical Computing).

## RESULTS

### Subject characteristics

Thirteen patients with AN (20.5±1.7 years) and thirteen HC (23.8±3.3 years) were enrolled in this study. The key characteristics of the study groups are shown in Table. As expected, patients in the AN group had significantly lower BMI (16.7±1.2 kg/m^2^ vs 22.8±1.9 kg/m^2^; p<0.0001), body fat mass (20.2±7.5 % vs 33.6±5.4 %; p<0.0001), systolic blood pressure (102.6±6.9 mmHg vs 113.5±10.7 mmHg; p=0.0066) and diastolic blood pressure (64.8±4.7 mmHg vs 76.8±9.1 mmHg; p=0.0005) as compared to the controls .Whole body insulin sensitivity M-value measured with hyperinsulinemic-euglycemic clamp per body weight and minute was significantly higher in AN than HC (40.1±13.4 µmol/(kg*min) vs 28.4±10.0 µmol/(kg*min); p=0.0197). All subjects underwent both [^11^C]carfentanil and [^18^F]FDG brain PET scans except for one AN who did not participate in the [^18^F]FDG PET scan.

### MOR availability, BGU, and BOLD signal

MOR availability from caudate (p=0.0192) and putamen (p=0.0264) was higher in AN than HC, and those from nucleus accumbens (NAcc) (p=0.0504) and thalamus (p=0.0850) showed the higher trend in patients with AN (Figure 1). Full-volume analysis revealed a consistent finding that shows higher MOR availability of caudate, putamen, NAcc, thalamus, and orbitofrontal cortex in AN (Figure 2). However, in [^18^F]FDG PET scans, BGU was not significantly different between AN and HC in both ROI analysis and voxel-wise analysis. MOR availability and BGU were negatively correlated in caudate (r=-0.4680; p=0.0183), NAcc (r=-0.4310; p=0.0315) and thalamus (r=-0.4094; p=0.0421) and showed the trend of negative association in putamen (r=-0.3880; p=0.0553) (Figure 3). In a subgroup analysis, MOR availability and BGU were negatively correlated in caudate (r=-0.6153; p=0.0332), NAcc (r=-0.6801; p=0.0149) and thalamus (r=-0.6173; p=0.0325) of AN, not in those of HC (Supplementary figure). In addition, there was no difference of hemodynamic brain activity of viewing palatable versus non-palatable foods between patients with AN and HC.

**Figure 1.**
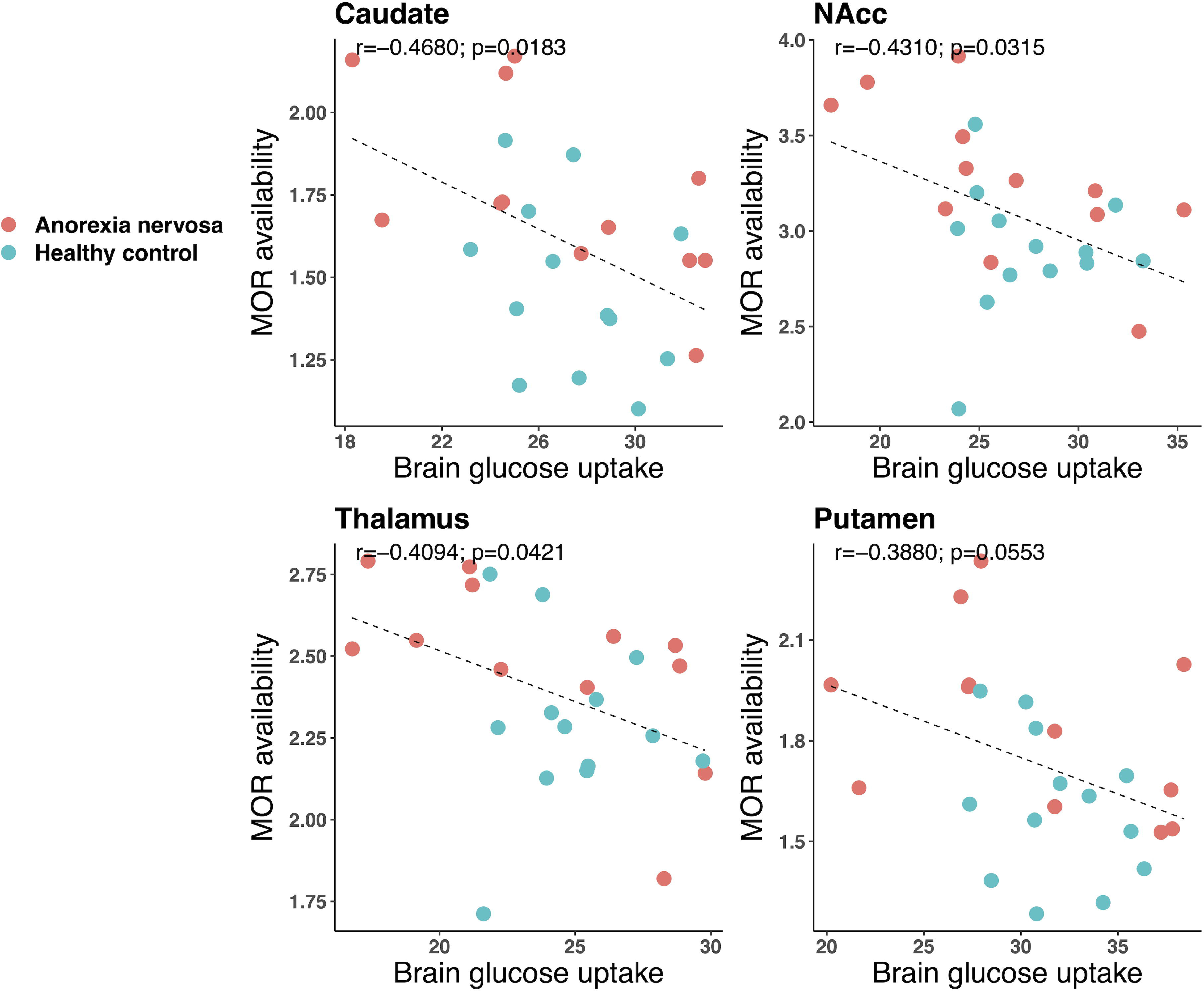
Comparison of MOR availability (BP_ND_) between anorexia nervosa and healthy controls

**Figure 2.**
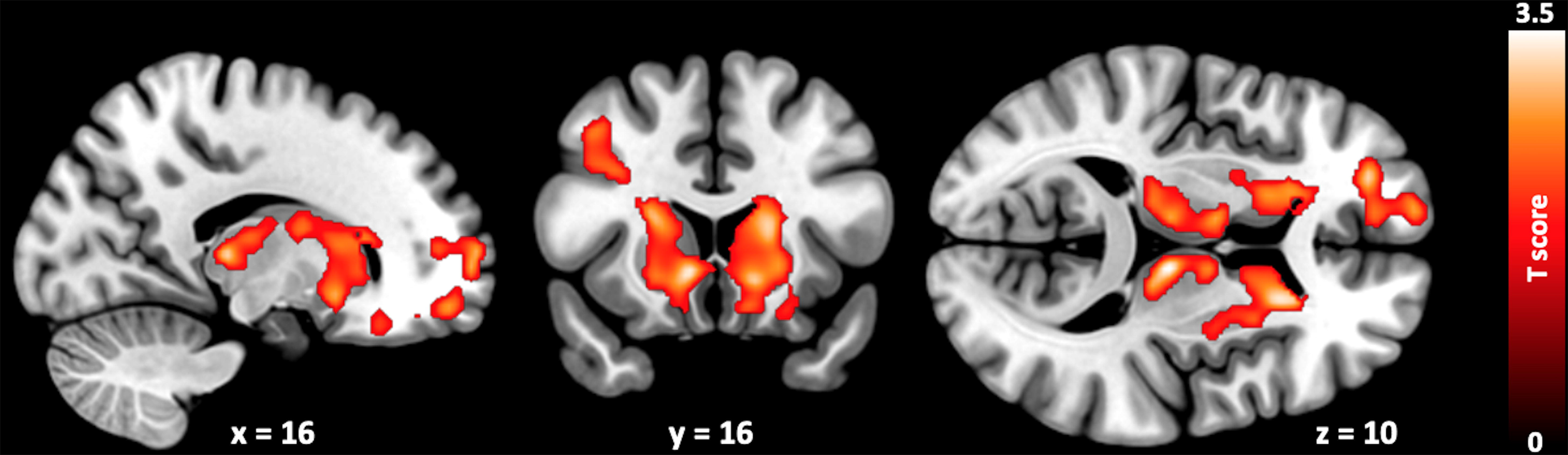
Brain regions showing higher MOR availability (BP_ND_) in anorexia nervosa versus healthy controls (FDR corrected p<0.05)

**Figure 3.**
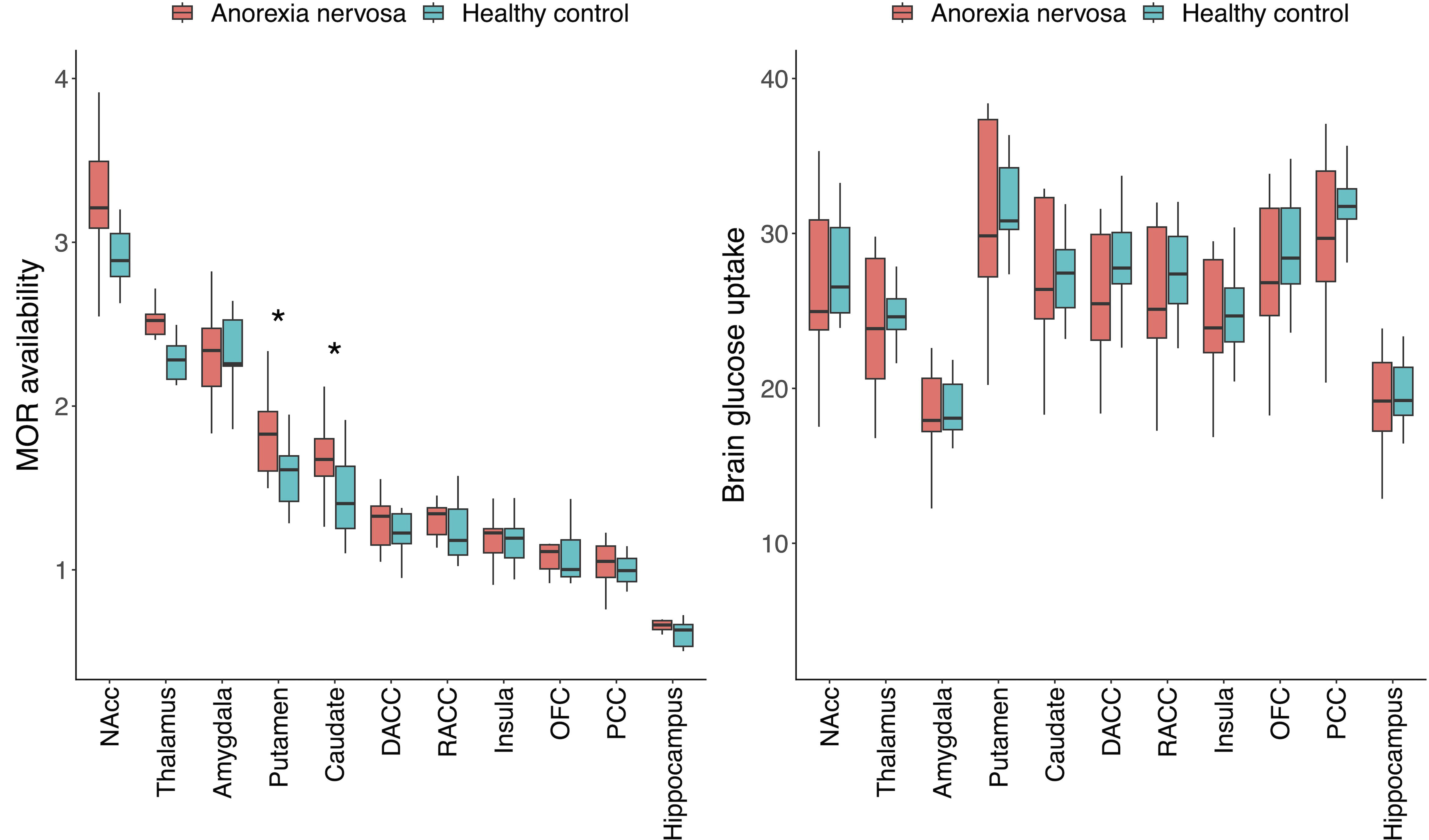
Correlation between MOR availability (BP_ND_) and BGU (μmol/min/100g) in anorexia nervosa and healthy controls

## DISCUSSION

Our main finding is that MOR availability in patients with AN was higher in brain regions implicated in reward processing, while BGU was not different between patients with AN and HC. Negative correlation between MOR availability and BGU was observed in caudate, NAcc, and thalamus. The effect of upregulated MOR availability is a mirror image of the downregulated MOR system in obesity ^13, 42, 43^. Studies have also found that upregulation of MORs occurs following weight loss ^42, 43^. Altogether these data show how cerebral glucose metabolism is maintained in typical levels even under severe undernutrition, whereas central MOR systems adapt to the peripheral dysregulation, potentially contributing to the mood changes in patients with AN.

### Upregulated Mu-Opioid Receptor Availability in AN

AN is a severe psychiatric disorder characterized by an extremely low body weight, fear to gain weight and a distorted body image with comorbid mood disorders, major depressive disorders, anxiety disorders, obsessive-compulsive disorders, autism and attention-deficit hyperactivity disorder^3^. The etiology of AN is complicated and involves genetic, neurobiological, psychological and environmental factors ^2^. Some patients with AN start losing the weight voluntarily and after several months, they have a positive experience with losing the weight, however, facing a difficultly to eat normally, causing discomfort and anxiety ^2^. As they have amplified negative experiences when trying to eat normally, the vicious cycle of AN is established ^2^. The treatment of AN includes medical care such as monitoring of vital signs and hydration, diet and psychotherapy to restore a healthy weight ^2^. However, the prognosis of AN is still poor with the highest mortality rate among all psychiatric disorders ^1^. In addition, post-treatment relapse rates exceed 30% ^44^. Thus, central molecular pathways contributing to feeding, mood, and reward would be linked with AN.

In line with this hypothesis, we found that central MOR upregulation was observed in patients with AN. The endogenous opioid system and particularly MORs are associated with both homeostatic and hedonic control of eating behavior ^18^ as well as mood regulation ^45^. Opioid antagonists decrease food intake and opioid receptor agonists stimulate food intake and the hedonic responses to palatable foods ^14^. Also, the consumption of both palatable and nonpalatable meals triggers the release of endogenous opioid ^15^. Individual variation in MOR is also associated with anticipatory reward responses, and lowered MOR availability is linked with stronger BOLD fMRI responses to appetizing foods in frontal and cingulate cortices, striatum and amygdala of the reward and emotion circuit ^26^. In the present study, upregulated MOR availability in patients with AN was observed in brain regions implicated in reward processing, which might be associated with reduced pleasure in eating behavior, a refusal to eat and this would lead to a vicious cycle of AN. This result is also in line with previous studies in obesity. Our group has previously shown that MOR availability is higher in nonobese subjects than morbidly obese subjects (22.7±2.9 kg/m^2^ vs 41.9±3.9 kg/m^2^) ^13^, and that was higher in patients with AN than HC (16.7±1.2 kg/m^2^ vs 22.8±1.9 kg/m^2^) in this study. Thus, it is possible that the MOR availability scales with BMI with highest availability in patients with AN and lowest in morbidly obese subjects, with HC in between. However, explanations based on reward-based versus metabolically driven alterations are not mutually exclusive, and cannot be dissociated in the present cross-sectional study.

Prior studies have found no difference of striatal dopamine receptor availability between patients with AN and HC ^9^, however, there is a possibility of MOR-induced disruption of the reward pathway through dopamine system in AN, which was reported in morbid obesity ^46^. One previous study with [^11^C]diprenorphine, a non-selective opioid receptor radiopharmaceutical, compared MOR availability between patients with AN and HC ^47^. Interestingly, opioid receptor availability was lower (rather than higher as in our study) in anterior cingulate cortex, insula, frontal, temporal and parietal lobes, suggesting the potential involvement of reward circuit modulation and anxiety in AN ^47^.

In addition to MORs, alteration of other neurotransmitter systems have been linked with AN. Increased serotonin 1A receptor of temporal, frontal lobes and amygdala ^6^ and decreased serotonin 2A receptor of frontal, parietal and occipital lobes ^7^ in patients with AN were reported after comparison with HC. Also, cannabinoid 1 receptor was increased in both cortical and subcortical areas of AN ^8^.

Involvement of serotonin and cannabinoid systems in AN is not surprising as both have an important role in eating behavior regarding reward pathway. Also, histamine 1 receptor of limbic system, particularly amygdala was significantly higher in AN with a possibility of the involvement of histamine system in psychological symptoms such as emotional changes to food and body image as well as eating behavior ^48^. However, there was no significant difference of dopamine receptor between patients with AN and HC ^9^, although mesolimbic pathway in AN works differently during reward learning and responding ^10^.

### Brain glucose uptake

In addition to MOR availability, we contrasted BGU between patients with AN and HC. Although there was no difference in BGU between patients with AN and HC, a negative correlation of M value, a measure of insulin sensitivity, with BGU was observed, consistent with previous findings^49^. Global cerebral hypometabolism was observed in patients with AN using dynamic acquisition of [^18^F]FDG PET without hyperinsulinemic-euglycemic clamp ^11^. This altered brain glucose metabolism may be a consequence of low body weight and prolonged starvation state induced acidoketosis ^50^ or a morphologic factor such as enlargement of cortical sulci and ventricles ^51^ or changes in the permeability of blood vessels ^11^. However, another study with static acquisition of [^18^F]FDG PET, hypermetabolism of frontal lobe, hippocampus, amygdala, insula and hypometabolism of parietal lobe was reported ^12^. The method of [^18^F]FDG PET acquisition might affect the result of BGU in AN and HC and, currently, [^18^F]FDG PET during hyperinsulinemic euglycemic clamp is the gold standard for measuring regional tissue glucose uptake rates ^52^. A negative correlation between MOR availability and BGU was observed in the same brain regions that showed the higher MOR availability in AN. These data show that BGU is maintained despite the overall poor metabolic states of patients with AN, could indicate that the brains have to be kept up and running at all costs. In addition, there is a possibility of altered MOR leading to the constant BGU state by the interaction between MOR and BGU in patients with AN.

### Strengths and Limitations

Our study has provided deep phenotypical evaluation of patients with AN and matched HC. The combination for the first time of two PET tracers is also a considerable strength. Our study also has limitations. First, as AN is more predominant in females, we included only females with AN in this study. Therefore, the results may not be applicable in males. Second, as patients with AN may be particularly sensitive to questions regarding their eating behavior, we did not perform a questionnaire of eating behavior on participants so we could not link the changes in MOR availability and BGU with their eating behaviors. Third, the study was relatively small, yet despite this we observed robust differences in MOR availability. Finally, higher MOR availability from [^11^C]carfentanil PET BP_ND_ may reflect either an increased number of receptor proteins or an increased affinity to bind this radioligand agonists and it remains unclear whether this change in opioid system is the cause or the result of AN.

AN is associated with higher MOR availability in the brain regions implicated in reward processing, while BGU remains unaltered. Therefore, the endogenous opioid system might be one of the key components underlying AN and concomitant alterations in food intake and mood. The better understanding of the pathophysiology of AN could eventually lead to the development of new treatments for AN.

## Data Availability

All data produced in the present study are available upon reasonable request to corresponding authors

## ACKNOWLEDGEMENTS

The study was supported by National Research Foundation of Korea (KP: 2020R1F1A1054201), Sigrid Juselius Foundation (LN), Gyllenberg’s stifftelse (LN) and Academy of Finland (LN: 294897 and 332225).

