## supplementary figures for "Anorexia nervosa is associated with higher brain mu-opioid receptor availability"

**Supplementary information**

Correlation between MOR availability (BP_ND_) and BGU (μmol/min/100g) in subgroups of anorexia nervosa and healthy controls. MOR availability (BP_ND_) and BGU (μmol/min/100g) were negatively correlated in caudate, NAcc and thalamus of anorexia nervosa, not those of healthy controls.

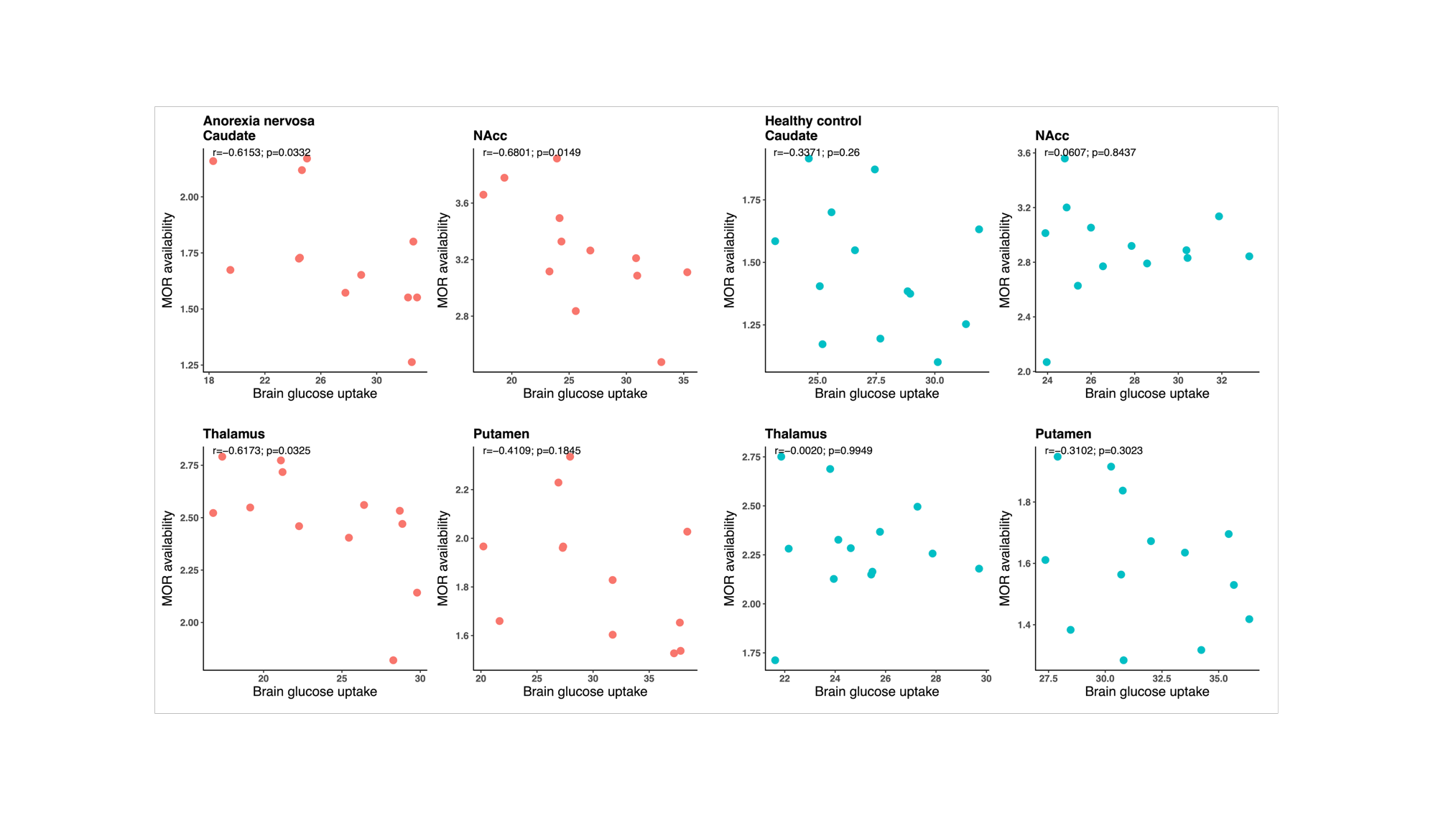
